# Performance of At-Home Self-Collected Saliva and Nasal-Oropharyngeal Swabs in the Surveillance of COVID-19

**DOI:** 10.1101/2020.10.23.20218487

**Authors:** Paulo H. Braz-Silva, Ana C. Mamana, Camila M. Romano, Alvina C. Felix, Anderson V. de Paula, Noeli E. Fereira, Lewis F. Buss, Tania R. Tozetto-Mendoza, Rafael A. V. Caixeta, Fabio E. Leal, Regina M. Z. Grespan, João C. S. Bizário, Andrea B. C. Ferraz, Dipak Sapkota, Simone Giannecchini, Kelvin K. To, Alain Doglio, Maria C. Mendes-Correa

## Abstract

SARS-CoV-2 quickly spread in the worldwide population by contact with oral and respiratory secretions of infected individuals, imposing social restrictions to control the infection. Massive testing is essential to breaking the chain of COVID-19 transmission. The aim of this study was to compare the performance of at-home self-collected samples - saliva and combined nasal-oropharyngeal swabs (NOP) - for SARS-CoV-2 detection in a telemedicine platform for COVID-19 surveillance. We analyzed 201 patients who met the criteria of suspected COVID-19. NOP sampling were combined (nostrils and oropharynx) and saliva collected using a cotton pad device. Detection of SARS-COV-2 was performed by using the Altona RealStar® SARS-CoV-2 RT-PCR Kit 1.0. According to our data, there was an overall significant agreement (κ coefficient value of 0.58) between the performances of saliva and NOP. Assuming that positive results in either sample represent true infections, 70 patients positive for SARS-CoV-2 were identified, with 52/70 being positive in NOP and 55/70 in saliva. This corresponds to sensitivities of 74.2% (95% CI; 63.7% to 83.1%) for NOP and 78.6% (95% CI; 67.6% to 86.6%) for saliva. We also found a strong correlation (β-coefficients < 1) between the cycle threshold values in saliva and NOP. Ageusia was the only symptom associated with patients SARS-CoV-2 positive only in NOP (p=0.028). In conclusion, our data show the feasibility of using at-home self-collected samples (especially saliva), as an adequate alternative for SARS-CoV-2 detection. This new approach of testing can be useful to develop strategies for COVID-19 surveillance and for guiding public health decisions.

## INTRODUCTION

Rapid and accurate diagnostic tests are essential for controlling the SARS-CoV-2 pandemic. Nevertheless, biological specimen collection is an important logistic challenge to provide massive testing(1-3). The possibility to use self-collected samples for COVID-19 testing offers several advantages, especially to minimize the risk of exposing healthcare workers to the virus, since self-collection does not require direct involvement of trained personnel in the sample collection(4, 5). Recently publications have been showed a similar sensitivity between saliva samples and nasal swabs collected by health-workers and those collected by a patient for COVID-19 molecular diagnosis, thus providing an important background for the choice of this strategy for surveillance of COVID-19(6-8).

Saliva has been described as a good alternative for SARS-CoV-2 detection, showing additional advantages compared to swab collection(9-12). Saliva collection does not cause discomfort or nasal bleeding to patients, does not require swab collectors or personal protective equipment, which are currently in short-supply in the market(3). In addition, saliva allows examination of several biomarkers, which could be useful as molecular signatures for patient stratification regarding infection severity(10, 13, 14).

In a pandemic scenario, at-home self-collection of samples plays a key role in the surveillance and control of the infection by allowing the patient with clinical suspicion of COVID19 to have access to proper healthcare and quick isolation of the confirmed cases(4, 5). Recently published studies show that 80-85% of individuals infected with SARS-CoV-2 have few or no symptoms, while 15-20% develop more severe disease, often associated with advanced age or other co-morbidities(15).

The Corona São Caetano program is a primary care initiative providing specific home care to all residents of São Caetano do Sul, state of São Paulo, Brazil. This program started in April 2020 due to the increasing number of COVID-19 cases in the country. Self-collection of nasal-oropharyngeal swabs has been used to obtain samples for diagnosis since the beginning of the epidemics, with excellent results, as hospitalization rate was less than 3% among the patients enrolled in the program(16).

The aim of this study was to compare the performance of two different at-home self-collected samples - saliva and combined nasal-oropharyngeal swabs (NOP) - for COVID-19 molecular diagnosis in the community patients outside the healthcare facilities.

## PATIENTS AND METHODS

The present study was developed in a telemedicine platform for COVID-19 surveillance called “Corona São Caetano”. Residents of the municipality aged 12 years or older who had suspected symptoms were encouraged to contact the program via website or by phone. They were invited to complete a screening questionnaire including socio-demographic data, information on type, onset and duration of the symptoms.

In the last two weeks of May 2020, a serie of 201 consecutive patients participating to the program were included in the present study. The patients met the defined criteria of suspected COVID-19 (i.e., having at least two of the following symptoms: fever, cough, sore throat, coryza or change in/loss of smell [anosmia]; or one of these symptoms plus at least two other symptoms consistent with COVID-19) were further evaluated. These patients were then called by a healthcare professional in order to complete a risk assessment. All pregnant women and patients meeting pre-defined screening criteria for severe disease were advised to attend a hospital service. All the other patients were offered a home visit for self-collection of saliva and NOP samples.

Patients testing RT-PCR positive for SARS-CoV-2 were followed up for 14 days (a maximum of 7 phone calls) after completion of their initial questionnaire, whereas those testing negative were followed up in the primary healthcare program. The patients were asked to contact the platform for a new consultation if they developed new symptoms.

This study was conducted according to ethical standards defined by institutional and national research ethics committees and the Helsinki Declaration of 1964, including subsequent amendments or comparable ethical standards, and approved by the Clinics Hospital Research Ethics Committee of the University of São Paulo School of Medicine under protocol number 3.979.632. Informed consent was obtained from all the individuals enrolled in this study. This study was designed conform to STROBE Guidelines.

### Sample Collection

NOP sampling were combined (both nostrils and oropharynx) using commercial flocked swabs with plastic applicators (Goodwood medical care Ltd., Jinzhou, China). Saliva samples were collected using a cotton pad device - Salivette™ (SARSTEDT AG & CO. KG, Nümbrecht, Germany). In order to provide guidance on self-collection procedures, a link to an instructional video was sent to each participant before the home visit. Briefly, patients were instructed to use the swabs in both nostrils and posterior region of the mouth and put both swabs into a tube containing saline solution. For saliva collection, they were instructed to chew carefully a cotton pad for one minute and put it into a specific tube. The samples were collected during the morning hours and the participants were instructed to avoid eating, drinking or toothbrushing at least one hour before the saliva collection. In accordance with the Corona São Caetano Program procedures, samples were immediately put in a cool box (2-8°C) and stored at 4°C in a fridge until shipment to the lab by a specialized carrier in the afternoon the same day.

### RNA Extraction and Real Time PCR

To recover the saliva from the devices, the tubes were centrifuged at 5,000g for 5 minutes. Total nucleic acid was extracted from 200 μl of the saline solution containing NOP and recovered saliva by using the NucliSENS EasyMag (BioMérieux, Durham, North Carolina, USA) automated DNA/RNA extraction platform.

Detection of SARS-COV-2 RNA was performed by using the Altona RealStar® SARS-CoV-2 RT-PCR Kit 1.0 (altona Diagnostics GmbH, Hamburg, Germany) which employs a B-COV specific probe directed to gene E and a SARS-COV-2-specific probe directed to gene S. Results were considered positive when one or both genes were amplified with a cycle threshold (Ct) < 40.

### Statistical Analysis

Cohen’s kappa coefficient (κ) was used to measure agreement between RT-PCR-based detection of SARS-COV-2 in saliva and NOP swabs. The sensitivity of each method was calculated assuming that positive cases in either sample type represented true positives, with 95% confidence intervals being calculated by using the exact method.

Next, we defined four analytic groups as follows: NOP-/saliva- (G1); NOP- /saliva+ (G2); NOP+/saliva- (G3); NOP+/saliva+ (G4). In order to identify clinical features associated with positivity in NOP and saliva, we specifically compared groups G2 to G4, G3 to G4, and G2 to G3. We used chi-squared test and Wilcoxon’s rank-sum test to compare clinical features (i.e. age, gender, symptoms and onset of illness) between the patients in these groups.

We then explored the relationship between RT-PCR cycle threshold (Ct) and sample type. We first analyzed group G4 (NOP+/saliva+) and assessed the association between Ct values by using the simple linear regression. Differences in the distribution of Ct values using NOP and saliva were assessed with paired Wilcoxon’s rank-sum test. We also assessed the association between time from symptom onset and collection of saliva and NOP by using the simple linear regression. Statistical significance was set at 0.05. All analyses were performed by using the R Statistical Software, version 3.6.3.

## RESULTS

For the current study, 201 consecutive patients participating to the Corona São Caetano program and who met the suspected COVID-19 case definition were included. RT-PCR-based COVID-19 testing was performed in samples from NOP and saliva and results are shown in Table 1.

**Table 1.** Comparison of SARS-CoV-2 RNA status in saliva and NOP samples in 201 patients undergoing testing for COVID-19.

| NOP | Saliva |  |
| --- | --- | --- |
|  | Negative | Positive |
| Negative | 131 | 18 |
| Positive | 15 | 37 |

Overall, 16,4 % (33/201) of the results were discordant giving a moderate agreement between the both sampling methods with a Cohen’s kappa coefficient of 0.58. Overall, assuming that positive test results in either sample type represent true infections around 35% of patients (n =70) were identified to be positive with COVID-19, while 26 % (n = 52) and 27 % (n = 55) were positive based on NOP or saliva detection, respectively. This corresponds to sensitivities of 74.2% (95% CI; 63.7% to 83.1%) for NOP and 78.6% (95% CI; 67.6% to 86.6%) for saliva samples. Of note, 9% (n=18) and 7% (n=15) resulted saliva or NOP single positive, respectively.

### Associations between Clinical Features and SARS-CoV-2 Positivity in Saliva and NOP samples using RNA RT-PCR

To investigate relationship between SARS-CoV-2 status in NOP and saliva and demographic and clinical features, patients were categorized in 4 groups (G1 to G4) (Table 2); NOP and saliva SARS-CoV-2 negative patients were grouped in G1 (NOP- /saliva-, n = 131); NOP-/saliva+ patients in G2 (n = 18); NOP+/saliva-in G3 (n = 15) and NOP+/saliva+ in G4 (n= 37). We didn’t found any significant relationship regarding the demographic data of patients in the different groups, although the sex ratio may be different in G2 as compared to G4. Also, no significant correlation was found regarding clinical symptoms, only ageusia appears to be more prevalent in patients with positivity of SARS-CoV-2 in NOP (p = 0.028). Interestingly, the delay between clinical symptoms onset and time of sample collection was significantly shorter (p < 0.05) in G2 as compared to G3 and G4, suggesting that patients with simultaneous SARS-CoV-2 positivity in NOP and saliva (G4-patients) were more prompt to quickly display clinical symptoms (Table 2).

**Table 2.**
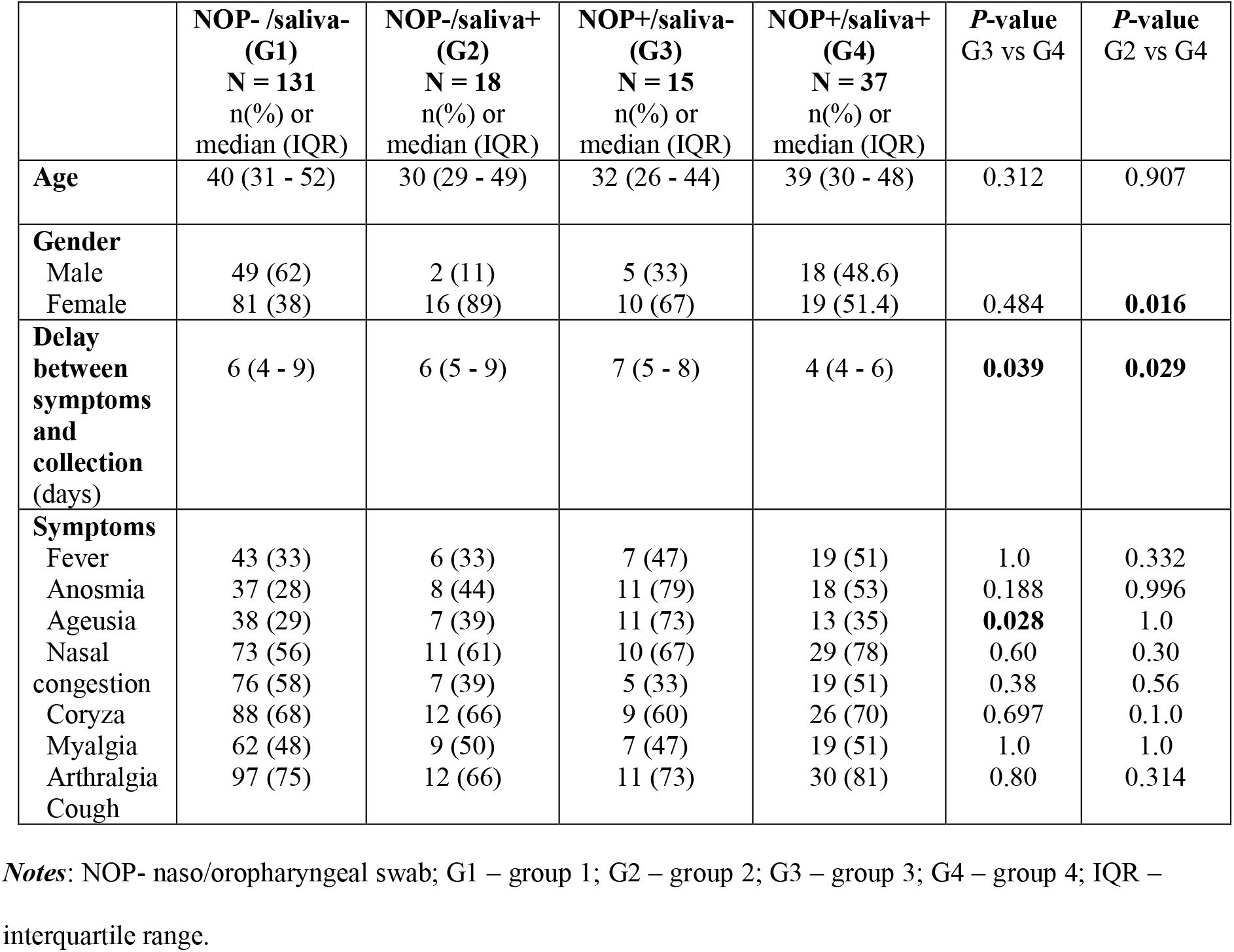
Demographic and clinical characteristics of 201 patients with suspected COVID-19 tested with RT-PCR in both saliva and NOP samples.

### RT-PCR Cycle Thresholds in Saliva and NOP Samples

In order to investigate the relationship between RT-PCR cycle threshold (Ct) and sample type (saliva or NOP), we first compared the Ct values from G4 (NOP+/saliva+, n =37) and assessed the association between Cts using simple linear regression. We found a strong correlation between the Ct values in saliva and naso/oropharyngeal samples (Figure 1). The coefficients of the regression lines (β) were 0.79 (p < 0.001) and 0.74 (p = 0.002) for E and S genes, respectively. A β-coefficient < 1 indicated that, in patients with SARS-CoV-2 positivity in NOP and saliva (G4), Cts tended to be higher in saliva then in NOP (also see Figure 2A).

**Figure 1.**
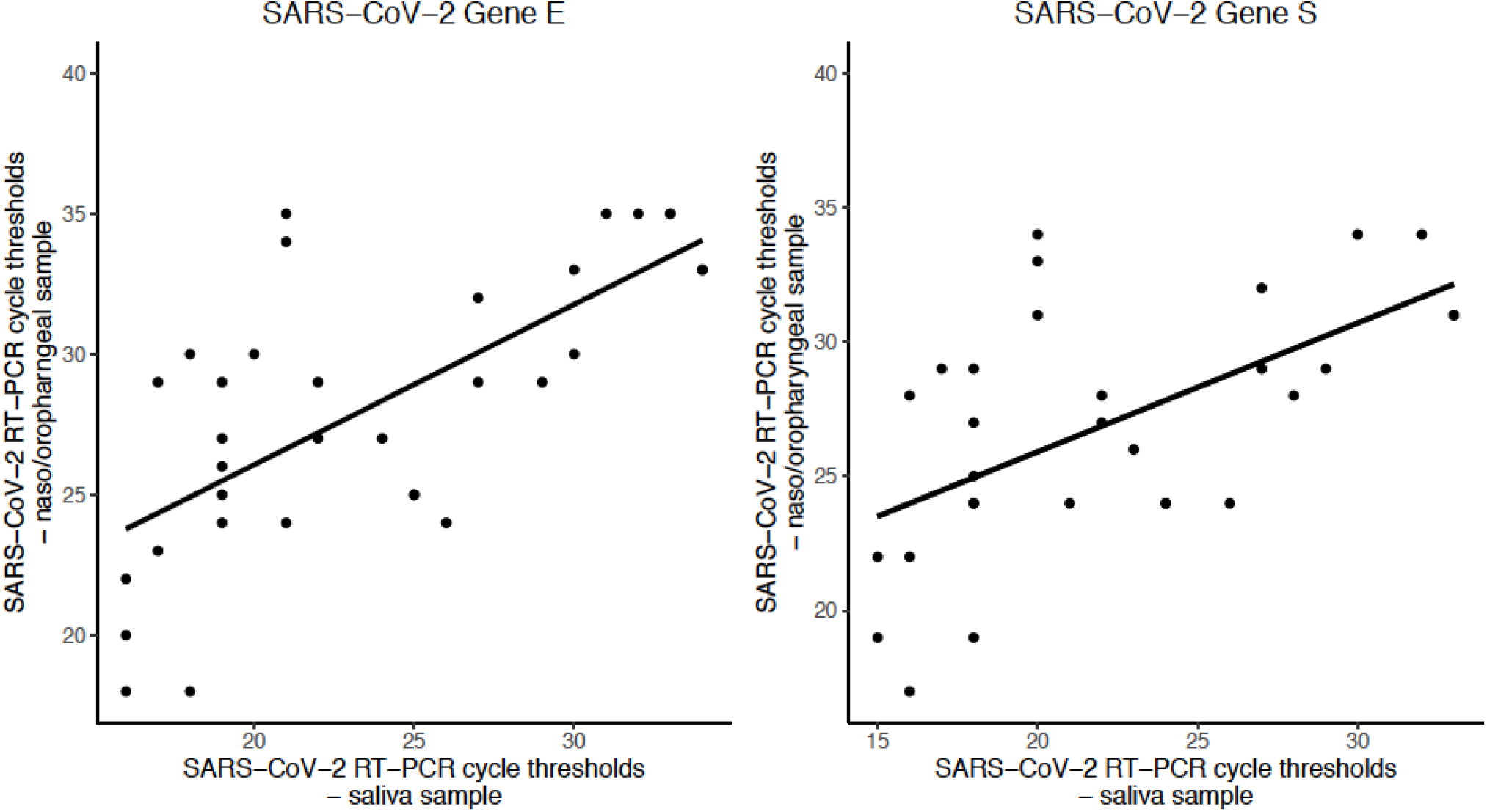
Comparison of RT-PCR cycle thresholds between naso-oropharyngeal and saliva samples in 37 patients with positive results in both samples. The coefficients of the regression lines are 0.79 (*P* < 0.001) for gene E and 0.74 (*P* = 0.002) for gene S.

**Figure 2.**
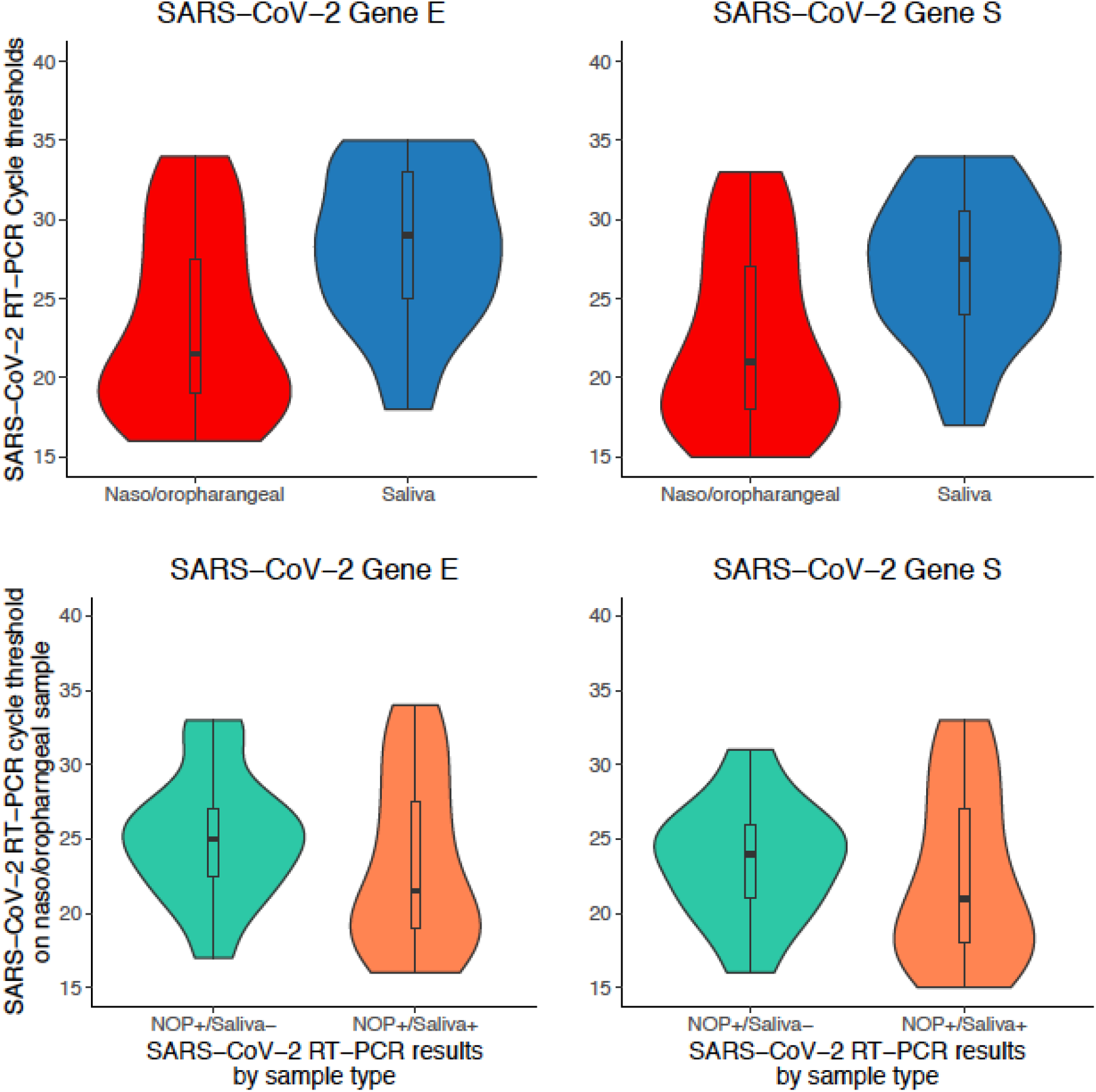
**A** - Violin plots showing the distribution of cycle thresholds in nasal-oropharyngeal swabs (NOP) and saliva samples for the two genes (E and S) amplified by RT-PCR. Boxplots shows median, interquartile range and range as standard. Analysis of the 37 patients with positive results in both sample types by comparing the distributions of cycle thresholds between NOP and saliva samples. Paired Wilcoxon’s rank-sum test was used, in which *P*-values were < 0.001 for genes E and S. **B** - Distribution of SARS-CoV-2 RT-PCR cycle thresholds in the 52 positive nasal-oropharyngeal swabs (NOP) samples stratified by RT-PCR results in saliva (NOP+/saliva- *versus* NOP+/saliva+). Boxplots shows median, interquartile range and range as standard. Distributions were compared by using paired Wilcoxon’s rank-sum test, in which *P*-values were 0.21 and 0.35 for genes E and S, respectively.

Next, we compared the Ct values between the groups G3 (NOP+/saliva-) and G4 (NOP+/saliva+). It was observed that the cycle threshold values were lower in patients positive in both NOP and saliva samples (median [IQR], 21.5 [19 - 27]) compared to those positive only in NOP samples (29 [25 - 33], *P* = 0.01, Wilcoxon’s rank-sum test) for gene E. Moreover, these results were for gene S (Figure 2B).

### Relationship between timing of sample collection and SARS-CoV-2 detection

Because timing of sample collection is a critical parameter of SARS-CoV-2 diagnosis, we further investigate the possible relationship between NOP and saliva SARS-CoV-2 detection and the delay between clinical symptom onset and timing of sample collection. Although Ct values tended to be higher at later periods from the onset of symptoms, this did not reach statistical significance (Figure 3).

**Figure 3.**
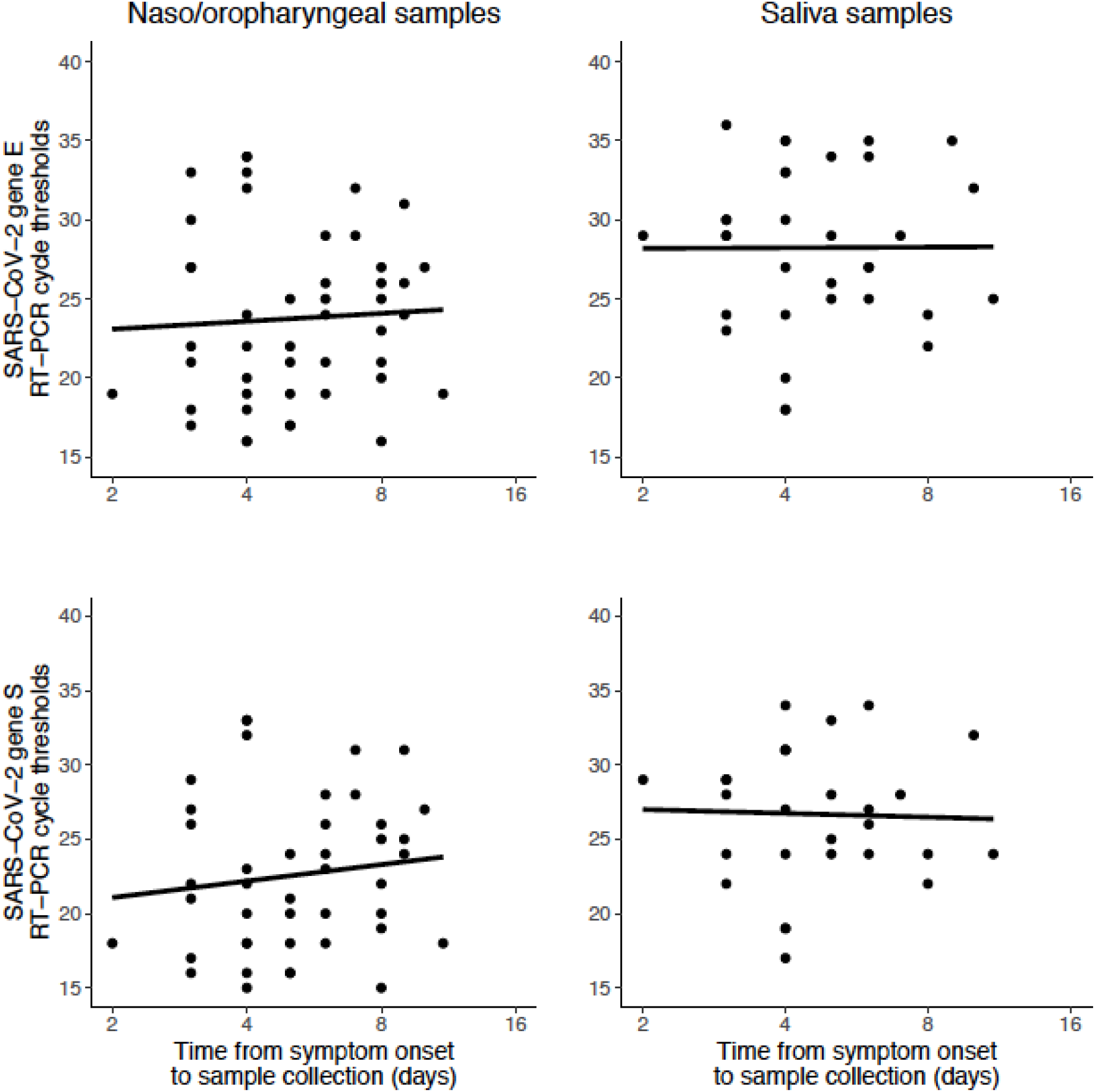
Relationship between illness course (i.e. time elapsed between symptom onset and sample collection) and cycle threshold values for nasal-oropharyngeal swabs (NOP) (left-hand panels) and saliva samples (right-hand panels). In the NOP samples, the regression coefficients for cycle threshold (delay of log_2_-days) for genes E and S were 0.5 (*P* = 0.72) and 1.1 (*P* = 0.42), respectively; the regression coefficients for saliva samples were 0.04 (*P* = 0.98) and −0.26 (*P* = 0.87) for genes E and S, respectively.

## DISCUSSION

We prospectively analyzed a cohort of patients with mild symptoms of COVID-19 to assess the diagnostic performance of at-home self-collection of combined naso-oropharyngeal swabs (NOP) and saliva samples.

According to our data, there was an overall significant agreement (κ coefficient value of 0.58) between the performances of saliva and NOP samples in the diagnosis of COVID-19. Assuming that positive results in either sample type represent true infections, a total of 70 patients positive for SARS-CoV-2 were identified, with 52/70 being positive in NOP and 55/70 in saliva. This corresponds to sensitivities of 74.2% (95% CI; 63.7% to 83.1%) for NOP and 78.6% (95% CI; 67.6% to 86.6%) for saliva samples. We also found a strong correlation (β-coefficients < 1) between the cycle threshold (Ct) values in saliva and NOP samples. However, the Ct values for the studied genes tended to be higher in saliva than in NOP samples.

The use of saliva to detect SARS-CoV-2 has been extensively analyzed by different authors, showing that saliva can be used as an alternative sample to nasopharyngeal swabs for COVID-19 molecular diagnosis(9, 10). The sensitivity found in their studies varied from 81% to 100%(11, 12, 17-21). The majority of the studies were conducted with hospitalized patients presenting more severe clinical forms of the disease, or patients attending a health care unit. It is important to highlight that even with a different population and different saliva collection (i.e. cotton pad device), we found similar sensitivity values.

To the best of our knowledge, the present study is the first one in the literature to prospectively examine the performance of at-home self-collected saliva and NOP, in a telemedicine based platform for COVID-19 surveillance. This is highly desirable in a pandemic scenario as it contributes to minimizing the risk of infection transmission in these settings. Additionally, it significantly reduces the workload burden of the healthcare units.

According to our results there was a moderate, but very significant, agreement (κ coefficient value of 0.58) between the performances of both sampling methods (NOP and saliva) in the diagnosis of COVID-19. This good overlap between both specimen types was in line with others studies where in general the agreement rate observed varied from 0.45 to 1(11, 22). In our study for around 16% of patients, RT-PCR results gave discordant results between both sampling methods. This variation in agreement may be associated with different factors, such as clinical characteristics of the population, diagnostic kits, saliva collection methods, among others. However, these inconsistent results are likely to be related to the fact the virus can reach oral and nasopharyngeal area with different kinetics being not always present at the same time in both sites as previously reported(23).

It has been postulated that there is a minimum of three different pathways for SARS-CoV-2 to reach the saliva: firstly, from the lower and upper respiratory tract; secondly, presence in the blood and gingival crevicular fluid; and thirdly, through salivary gland infection, with subsequent release of viral particles into the saliva via salivary ducts(10, 24, 25). It is believed that the highest viral concentration observed in saliva is derived from the respiratory tract(12). Therefore, the finding of viruses in saliva would be expected only in cases with a higher viral load, since the viral particles observed in the saliva also depends on the amount of viruses coming from the respiratory tract. However, the fact that in our study 9% of the samples were positive in saliva in absence of NOP positivity could be indicative that at least in a minimal part the virus comes from salivary origin.

In addition, based on the 37 patients positive in both saliva and NOP samples, we found a strong correlation between cycle threshold (Ct) values in saliva and those in NOP samples, indicating that Ct values tended to be higher in saliva than in NOP samples (β-coefficients < 1). When we compared the cases positive in both methods to those positive in NOP only, it was found that Ct values were lower in the first group. Th**i**s finding reinforces the idea that the viral load has to be higher (lower Ct) in order to be positive in the saliva. Clearly, according to our results, the viral load influenced the results. Only cases with a higher viral load (lower Ct) were positive in both methods, whereas cases with higher Ct values were positive in NOP samples only.

Some studies have compared the viral load between nasopharyngeal swab and saliva samples, showing a tendency for a higher viral load (or lower Cts) in nasopharyngeal swabs(10, 19). The severity of the cases included and the time elapsed between collection of material and onset of symptoms are essential information to interpret correctly these results, since the sensitivity of the diagnostic methods varies according to these variables.

When we compared the chance of identifying the virus in saliva and NOP samples in relation to the time interval between onset of symptoms and sample collection time, it was observed that the identification of the virus in both samples were associated with a shorter interval of time. These results stress the importance of early diagnosis of COVID-19, in which sample should be collected within the first days of symptoms, thus minimizing the loss of sensitivity of the molecular diagnosis(26). Different studies have analyzed the SARS-CoV-2 shedding in different biologic specimens, reporting that viral loads from upper respiratory tract samples peak within a week of symptom onset and follow a relatively consistent downward trajectory(27). Viral load in other biologic specimens, including saliva, follows the same trajectory(28). However, according to these studies, viral load does not seem to be as high as that observed in respiratory tract samples. Therefore, our findings are in line with these observations.

Chemosensory deficits associated with SARS-CoV-2 infection are quite frequent among patients with mild or moderate disease, considered a very early symptom. Interestingly, in our study ageusia was the only symptom statistically associated with patients SARS-CoV-2 positive only in NOP samples (G3; p=0.028). This results corroborate with the role of the neurotropic and neuro-invasive characteristics of coronaviruses in the pathogenesis of ageusia, more than a local infection of the gustatory buds(29).

Recent studies comparing samples collected by a specialized health-workers and self-collected by the patients for COVID-19 molecular diagnosis showed that both methods had similar sensitivity, which highlights the reliability of self-collection as a public health strategy for COVID-19 surveillance(6-8). Our results corroborate these findings as they showed that both self-collected samples had good sensitivity, especially the saliva, with 78.6% (95% CI; 67.6% to 86.6%).

The present study showed that self-collection of saliva and NOP for diagnosis of COVID-19 is feasible in the studied population. Given the similar sensitivities of saliva and NOP samples for detection of SARS-CoV-2 in patients with mild symptoms, it is expected that self-collection of either sample can be valuable in the surveillance of COVID-19 at a population level(30). Moreover by simplifying the procedure and, above all, avoiding the need for the patient to go to a specialized laboratory, this innovative approach can improve COVID-19 diagnosis, notably allowing the sample to be collected as soon as possible after appearance of the first symptoms. However, in this sense, the ease of collection and feasibility for examination of molecular biomarkers for disease stratification and prognosis justify the use of self-collected saliva as a preferred biological sample(2, 26).

As the main limitation of this study, we can cite the non-inclusion of asymptomatic individuals in the platform of COVID-19 surveillance.

In conclusion, our data show the possibility of using at-home self-collected samples (especially saliva), as an adequate alternative for SARS-CoV-2 detection. This new approach of testing can be useful to develop strategies for COVID-19 surveillance and for guiding public health decisions.

## Data Availability

The data that support the findings of this study are available from the corresponding author, Braz-Silva PH, upon reasonable request.

## Author Contributions

P.H. Braz-Silva, M. C. Mendes-Correa contributed to conception and study design, analysis and interpretation of data, drafted and critically revised the manuscript; A.C. Mamana, C. M. Romano, L. F. Buss contributed to study design, acquisition, analysis, and interpretation of data, drafted the manuscript; A. C. Felix, A. V. de Paula, N. E. Ferreira contributed to acquisition, analysis, and interpretation of data, critically revised the manuscript; T. R. Tozetto-Mendoza contributed to conception of the study, interpretation of data, critically revised the manuscript; R. A. V. Caixeta contributed to analysis and interpretation of data, drafted the manuscript; F. E. Leal, R. M. Z. Grespan, J. C. S. Bizário, A. B. C. Ferraz contributed to conception and study design, acquisition of data, critically revised the manuscript; D. Sapkota, S. Giannecchini, K. K. To, A. Doglio contributed to analysis and interpretation of data, drafted and critically revised the manuscript. All authors gave their final approval and agree to be accountable for all aspects of the work.

## Acknowledgements

This study was supported by the Internal funding from the Hospital das Clínicas of the University of São Paulo School of Medicine, São Paulo, Brazil. We would like to thank José Tadeu Sales for the language correction of the manuscript. The authors declare no potential conflict of interest with respect to the authorship and/or publication of this article.

